# COVID-19 Vaccine Uptake and its Determinants in Cameroon: A Systematic Review and Meta-analysis

**DOI:** 10.1101/2025.01.12.25320427

**Authors:** Fabrice Zobel Lekeumo Cheuyem, Adidja Amani, Chabeja Achangwa, Brian Ngongheh Ajong, Claude Axel Minkandi, Myriam Mathilde Mbia Kouda Zeh, Larissa Linda Eyenga Ntsek, Jacques Philippes Essomba, Rudy Chiozem Jiogue, Olivier Ndagijimana, Ndzi Etienne Nchanji, Celestin Danwang

## Abstract

**Background:** COVID-19 vaccination is crucial for mitigating the pandemic’s impact. However, vaccine hesitancy and access challenges have hindered global vaccination efforts. This meta-analysis aimed to estimate the pooled COVID-19 vaccine uptake proportion and identify associated determinants in Cameroon.

**Methods:** This review, conducted according to Preferred Reporting Items for Systematic Reviews and Meta-Analyses (PRISMA) guidelines, identified articles through searches of electronic databases, including PubMed, Scopus, Web of Science, ScienceDirect, and Google Scholar, as well as through gray literature. The search encompassed published and unpublished studies from 2021 to 2024 reporting on COVID-19 vaccine uptake and/or acceptance in Cameroon. Extracted data were compiled in a Microsoft Excel spreadsheet and analyzed using R statistical software (version 4.4.2). A random-effects model was employed when heterogeneity exceeded 50%. Publication bias was assessed using funnel plots, Egger’s test, and Begg’s test. Meta-regression was used to explore the influence of study characteristics.

**Results:** Twenty-two studies, encompassing 24,130 participants, were included. The pooled vaccine uptake proportion was 37.14% (95% CI: 29.24-45.05), with substantial heterogeneity (*I*² = 98.2%, *p*<0.001). Subgroup analyses revealed lower uptake among the general population (23.18%; 95% CI: 10.11-36.25) and in community settings (16.0%; 95% CI: 0.97-31.04) compared with healthcare workers (42.12%; 95% CI: 34.14-50.09). Younger age (OR = 0.53; 95% CI: 0.42-0.67) was inversely associated with vaccine uptake, while being in a partnership (OR = 1.59; 95% CI: 1.11-2.27) was positively associated. Higher levels of education (OR = 1.75; 95% CI: 1.56-1.97), urban residence (OR = 1.66; 95% CI: 1.21-2.29).

**Conclusion:** This meta-analysis revealed a suboptimal pooled COVID-19 vaccine uptake required to ensure a herd immunity. The results of this meta-analysis underline the crucial need to step up efforts to improve vaccination coverage, particularly among the most vulnerable populations. Identifying and addressing the factors underlying this low coverage is imperative if public health objectives are to be met. Public health interventions should be tailored to address the specific concerns and needs of different age groups and marital statuses.

## Background

Since the initial report of a case in Wuhan, China [1], Coronavirus Disease 2019 (COVID-19) has disseminated rapidly across the globe, subsequently becoming a significant public health challenge [2]. Vaccination has historically played a crucial role in enhancing global public health outcomes, contributing to increased life expectancy and representing a highly cost-effective intervention for disease prevention [3].

During the pandemic an effective vaccine utilization was imperative for mitigating the associated social and economic burden and for establishing a viable exit strategy from the COVID-19 pandemic [1]. In this regard, global analysis estimated that COVID-19 vaccination during 2020-2024 saved 2.5 million lives, or 15 million life-years corresponding to approximately 1% of total global mortality during that period [4]. A study reported that COVID-19 vaccines reduced deaths by 59% overall representing approximately 1.6 million lives saved in Europe [5].

Cameroon’s COVID-19 vaccination outbreak began after the first confirmed case was reported on March 6, 2020. The government promptly introduced vaccination as part of its preventive measures. [6]. The country’s health authorities, led by the National Immunization Technical Advisory Groups and the Scientific Advice for Public Health Emergencies, approved four vaccines against COVID-19 [7]. However, the vaccination coverage remained relatively low. As of November 18, 2022, approximately 5% of the eligible population had been vaccinated, putting the nation far behind the worldwide goal of obtaining 70% vaccination coverage by the end of the year [8,9]. The Janssen vaccine was the most administered and at that period, notable gender and geographical disparities in vaccine uptake were observed in the country [6,10].

Despite the availability of various vaccines against COVID-19, ensuring equitable access for everyone remained a significant challenge [2]. The efficacy of the COVID-19 vaccines in controlling disease transmission and save lives was contingent upon achieving sufficient vaccine coverage [7]. However, concerns regarding potential adverse events and uncertainties about vaccine effectiveness have contributed to COVID-19 vaccine hesitancy [7,10]. Vaccine hesitancy is recognized as one of the top ten leading threats to global health. A distorted perception of disease risk, inadequate knowledge regarding vaccines, apprehension about adverse reactions, proliferation of misinformation and unsubstantiated claims are some of the key elements contributing to this public health problem [11]. Vaccine hesitancy adversely affected the vaccine uptake and impeded efforts to control the pandemic [12]. COVID-19 vaccine hesitancy is associated with a range of factors, including sociodemographic factors (female gender, younger age, nursing profession, lack of prior influenza vaccination), concerns about vaccine safety and efficacy, and mistrust of government and institutions [6,10,13].

Although the COVID-19 public health emergency of international concern was declared over in May 2023, current WHO recommendations emphasize the continued need to vaccinate priority populations at highest risk for severe COVID-19 disease and mortality. These recommendations also advocate for building more sustainable programs by integrating COVID-19 vaccination into primary health care, strengthening immunization across the life course, and improving pandemic preparedness [14].

The optimal implementation of these recommendations at national level required monitoring of evidence-based data to inform decision-making. In this regard, several studies conducted in Cameroon have assessed COVID-19 vaccine uptake and associated factors. These reports underscore variable degree of vaccine acceptance and uptake. To the best of our knowledge no previous meta-analyses of these COVID-19 vaccine indicators have been conducted in Cameroon. Therefore, the present study aimed to determine the pooled estimate of vaccine uptake and determinants in Cameroon.

## Methods

### Study Design

This study was conducted to assess the proportion of vaccine uptake and associated factors. The study results are reported based on the Preferred Reporting Items for Systematic Review and Meta-analysis (PRISMA) guidelines [15].

### Study Setting

Cameroon occupies a total land area of 472,650 km² and geographically divided into ten administrative regions: Centre, Littoral, Far North, North, Adamawa, North-West, South-West, West, East and South. Demographic data for 2023 indicate an estimated population of approximately 28.6 million. The nation has a dual capital structure: Yaoundé, located in the Central Region, serves as the designated political capital, while Douala, located in the Littoral Region, serves as the primary economic center driving national economic growth [16].

### Eligibility Criteria

This systematic review included all existing published and unpublished research documenting the COVID-19 vaccine uptake and willingness to accept the in Cameroon. Studies lacking clearly defined outcome variables were excluded. Duplicate articles were identified and subsequently removed prior data extraction. The review was restricted to articles published in English or French. No temporal restrictions on publication date were imposed, as there were no prior systematic reviews and meta-analyses investigating vaccine uptake in the country at the time of the search.

### Article Searching Strategy

A systematic search of electronic databases, including PubMed, Google Scholar, Scopus, and ScienceDirect, was performed to identify published studies. The search strategy included analysis of the text within the title and abstract of each study. A combination of keywords and Medical Subject Headings (MeSH) terms was employed, utilizing Boolean logic operators (“AND” and “OR”) to refine the search. The keywords and MeSH terms included “coronavirus OR COVID-19 AND vaccine AND acceptance OR willingness OR hesitancy OR uptake OR intention OR perception AND Cameroon.” To ensure a comprehensiveness, a manual search was conducted to identify additional published articles not indexed in electronic databases. Unpublished studies were sought at the University of Yaoundé I library. Furthermore, the reference lists of identified studies were screened to identify further relevant articles. The last search was conducted on November 15, 2024.

### Data Extraction

Data were extracted from all eligible articles using a predefined Microsoft Excel 2016 form to collect study characteristics. For the first outcome (vaccine uptake assessment), the data extraction checklist included the first author’s name, study year, region, study design, setting, study participants, sample size, reported vaccine uptake. For the secondary outcome (assessment of determinants), extracted data included the crude odds ratios and their corresponding confidence intervals. The natural logarithm of each odds ratio and its corresponding upper and lower limits were calculated based on the results reported in the original studies. Two authors independently assessed each article for relevance and quality. Discrepancies between reviewers were resolved through discussion with a third reviewer to achieve consensus.

### Data Quality Assessment

The quality of the included studies was assessed using the Joanna Briggs Institute (JBI) quality assessment tool for prevalence studies [17]. Nine criteria were employed to assess the risk of bias for each study. These criteria included appropriateness of the sampling frame, use of a suitable sampling technique, adequate sample size, description of study subjects and setting, sufficient data analysis, use of valid methods for identifying conditions, use of valid measurements for all participants, use of appropriate statistical analysis, and an adequate response rate (≥60%). Each criterion was scored as 1 (yes) or 0 (no or unclear). The risk of bias was categorized as low (5– 9), moderate (3–4), or high (0–2).

### Outcome Measurement

This systematic review and meta-analysis had two main outcomes. The primary outcome was COVID-19 vaccine uptake, estimated as the total number of individuals who received the COVID-19 vaccine divided by the total number of participants who responded to the question, expressed as a percentage. The secondary outcome was the determinants of willingness to accept the COVID-19 vaccine and actual vaccine uptake. These determinants were assessed using odds ratios calculated from binary outcomes reported in primary studies. The key factors identified through review of the primary articles were age (<30 vs. ≥30 years), sex (female vs. male), educational level (primary vs. none, secondary vs. none/primary, tertiary vs. none/primary), marital status (in a partnership vs. other), religion (other vs. Christian), professional group (other vs. doctor), employment status (unemployed vs. employed), residence (urban vs. rural), number of household members (<5 vs. ≥5), and past history of comorbidity (yes vs. no).

### Operational Definition

Vaccine uptake was defined as having received at least one dose of any COVID-19 vaccine approved in Cameroon [7]. Vaccine acceptance refers to the intention or willingness to receive the vaccine, not the actual administration (uptake) of the vaccine itself [12]. It was defined as a “yes” response to the question, “Will you accept COVID-19 vaccination if it was available?”.

### Statistical Analysis and Synthesis

Heterogeneity between studies was assessed using the *I*² statistic. Heterogeneity was then categorized as low (<25%), moderate (25-75%), and high (>75%). Subgroup analysis was performed for study year, region, setting, and type of participants enrolled. A random-effects model was employed when heterogeneity exceeded 50%. Meta-regression was performed to investigate whether study characteristics could explain the variability in results across studies. The examined study characteristics included the year the study was conducted (≤2022 or >2022), region (other Regions vs. Centre), setting (other settings, including online- and community vs. hospital-based), sample size (<300 and ≥300), study participant type (healthcare workers vs. general population), and sampling method (probabilistic vs. non-probabilistic). Only study variables with meaningful and practical categories were considered. Univariable and multivariable meta-regression models were used to assess whether vaccination uptake varied according to the selected explanatory variable categories. A *p*-value<0.05 was considered statistically significant. The “meta” package was used to perform analysis using R Statistics version 4.4.2 [18].

### Publication Bias and Sensitivity test

Publication bias was assessed visually using the funnel plot. A funnel plot displaying a symmetrical, inverted funnel shapes suggested the absence of publication bias. To further investigate potential publication bias, Egger’s linear regression and Begg’s rank correlation tests were performed, with a significance level of *p*<0.05. Sensitivity analysis was conducted by iteratively excluding one study at a time to explore the robustness of the findings.

## Results

A total of 1603 records were retrieved from the database search (n=1599) and from unpublished research studies (gray literature; n=4). After removing 167 duplicate records, 1436 records remained. Titles/abstracts, followed by full text articles were then screened for eligibility. Ultimately, 22 study reports met the eligibility criteria and were included in this systematic review and meta-analysis (Fig. 1).

### Selection of Studies

**Fig. 1.**
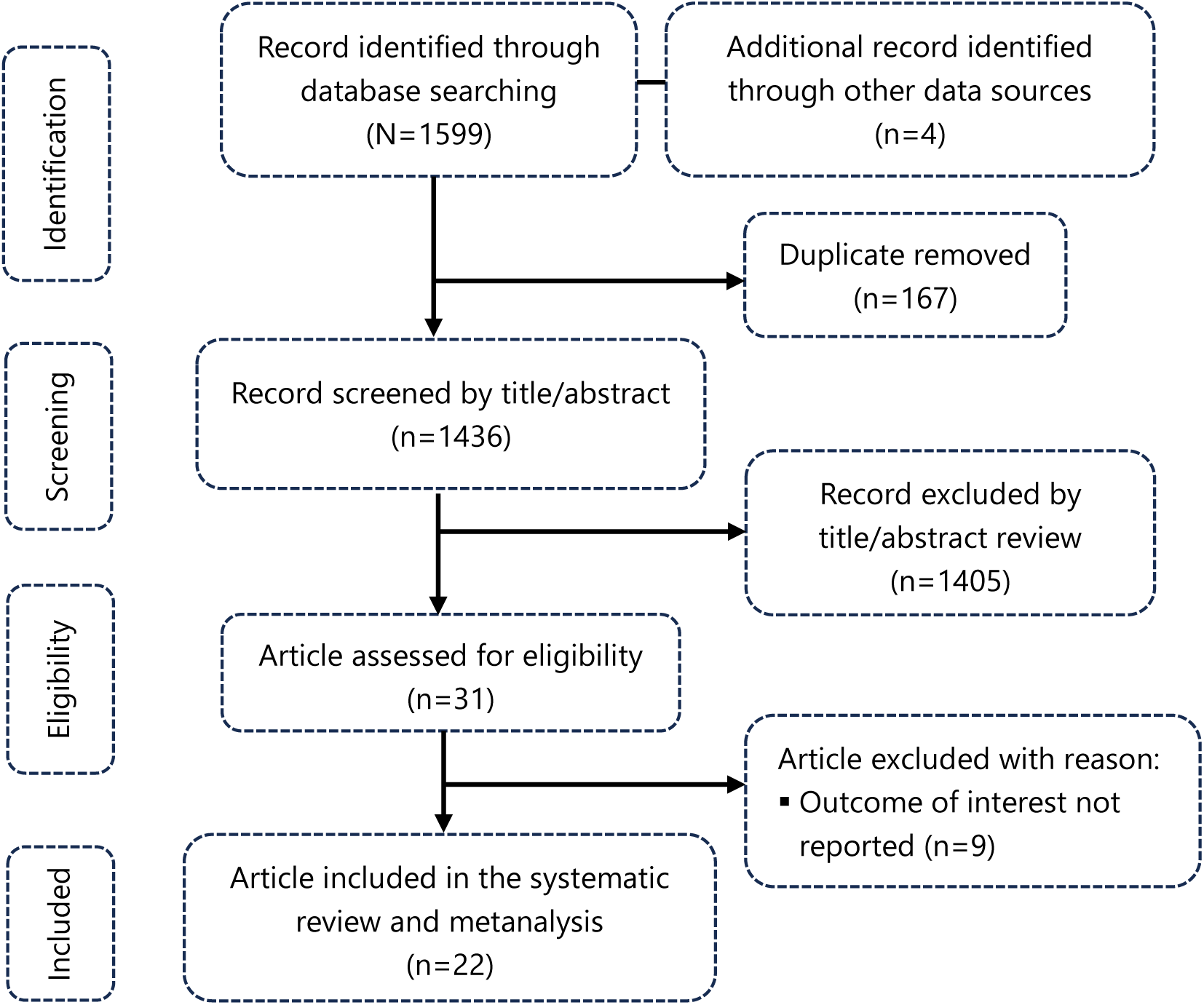
PRISMA diagram flow of studies included in the metanalysis

### Characteristic of Reports Included

A comprehensive analysis included 22 studies encompassing a total 24,130 participants. These studies, conducted between 2021 and 2024, involved both general population and healthcare workers in healthcare settings across the country and provided estimates of COVID-19 vaccine uptake and determinants of acceptance and uptake. All included studies employed a cross sectional design (Table 1).

**Table 1.**
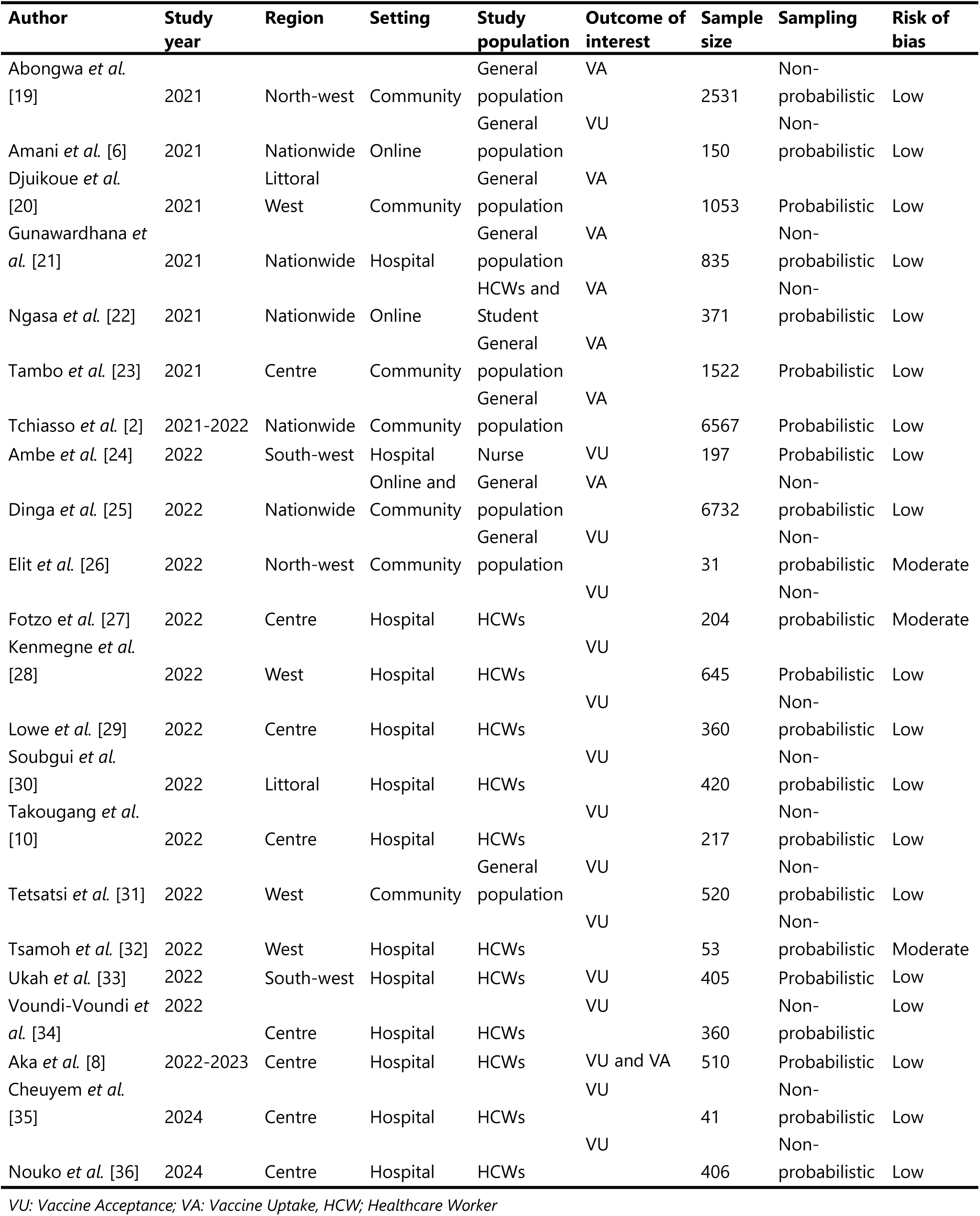
Characteristic of studies assessing compliance with the COVID-19 vaccine in Cameroon, 2021-2024.

### COVID-19 Vaccine Uptake

The overall pooled vaccine coverage was 37.14% (95% CI: 29.24-45.05); *I^2^*=98.2% with *p*˂0.001 (Fig. 2).

**Fig. 2.**
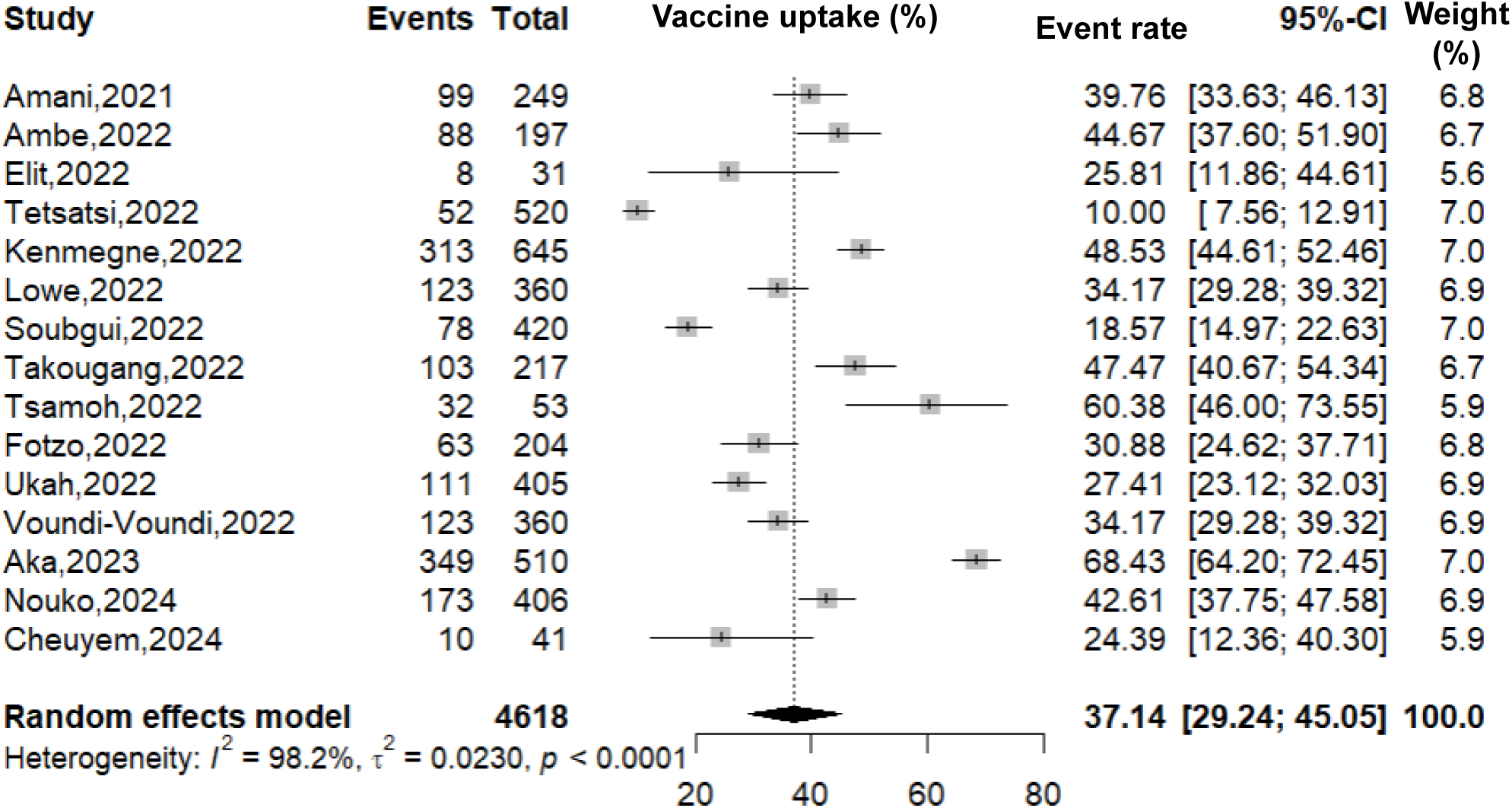
Pooled COVID-19 vaccination coverage in Cameroon, 2021-2024

The lowest pooled estimates of COVID-19 vaccine uptake were observed in studies conducted within communities (16.0%; 95% CI: 0.97-31.04, *n*=2 studies) and among general population (23.18%; 95% CI: 10.11-36.25, *n*=4 studies). Regarding geographical trends, with the exception of the North-west Region (which contributed only one study), the lowest coverage was observed in the South-west Region (35.82 %; 95% CI: 18.91-52.73, *n*=2 studies). The most recent pooled estimate (2024) was 34.57% (95% CI: 16.84-52.30, *n*=2 studies). The highest pooled estimate was observed among healthcare workers 42.12% (95% CI: 34.14-50.09, *n*=11 studies) (Fig. 3 and 4).

**Fig. 3.**
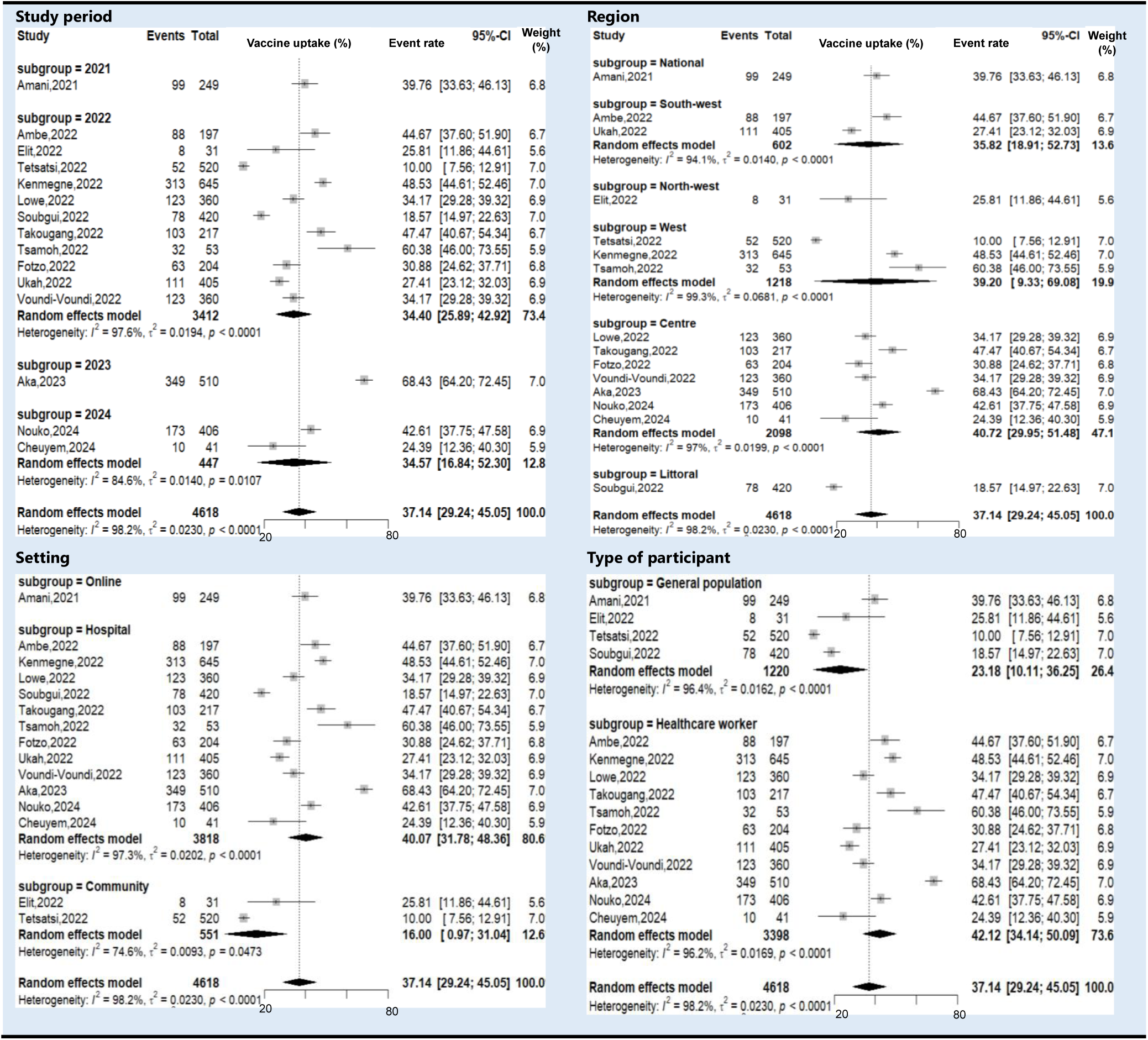
Subgroup analysis of the COVID-19 vaccine coverage in Cameroon, 2021-2024

**Fig. 4.**
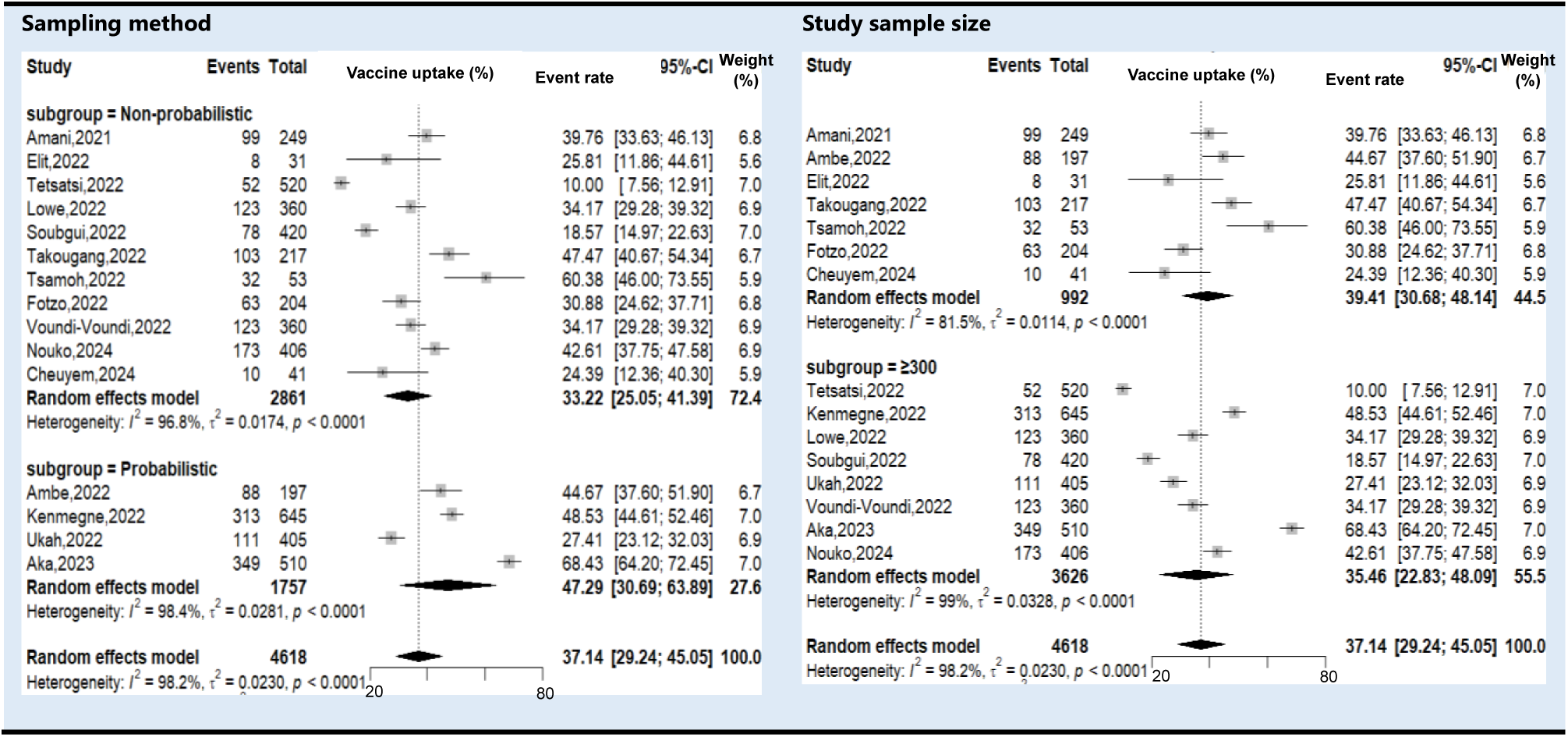
Subgroup estimates of the COVID-19 vaccine coverage in Cameroon, 2021-2024 *(a: by sampling methods; b: by study sample sizes)*

### Meta-Regression Analysis

Univariate analysis demonstrated that healthcare workers were significantly more compliant with the COVID-19 vaccine compared with the general population (β=0.9747; *p*=0.008). However, no parameter was significantly associated with the COVID-19 vaccine uptake at multivariate meta-regression analysis (Table 2).

**Table 2.** Multivariate metanalysis of COVID-19 vaccine uptake, 2021-2024.

| Category | Moderator | Univariate |  | Multivariate |  |
| --- | --- | --- | --- | --- | --- |
| | | coefficient ($\beta$) | p-value | coefficient ($\beta$) | p-value |
| Study year | ≤2022 vs. >2022 | -0.5279 | 0.271 | -0.3104 | 0.591 |
| Setting | Other<sup>1</sup> vs. Hospital | -0.8078 | 0.079 | 0.0346 | 0.973 |
| Region | Other<sup>2</sup> vs. Centre | -0.3613 | 0.352 | -0.0237 | 0.978 |
| Sampling | Probabilistic vs. Non-probabilistic | 0.6426 | 0.117 | -0.5085 | 0.429 |
| Sample size | <300 vs. ≥300 | 0.2381 | 0.549 | 0.4183 | 0.315 |
| Participant | Healthcare worker vs. General population | 0.9749 | 0.008 | 0.6581 | 0.502 |

<sup>1</sup>Online and/or within Community; <sup>2</sup>Multicenter and/or other specific Region

### Publication Bias and Sensitivity test Analysis

A large and almost symmetrical distribution of data points was observed in the funnel plot suggesting a low risk of publication bias. In addition, the Egger’s linear regression (*p*=0.287) and Begg’s rank correlation (*p*=0.217) tests confirmed the absence of statistically significant publication bias (Fig. 5).

**Fig. 5.**
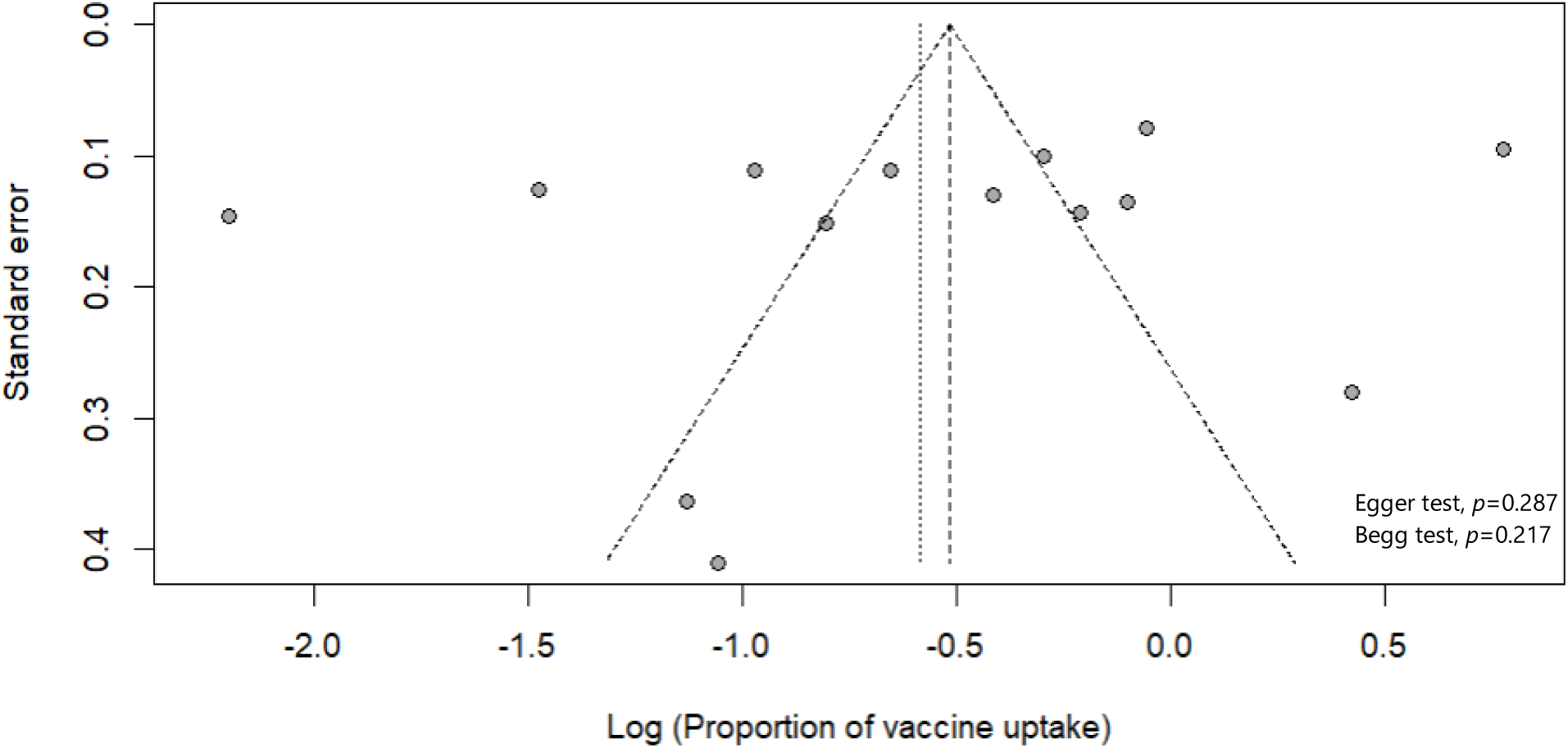
Funnel plot with pseudo 95% confidence limits and tests assessing the publication bias studies included

Sensitivity analysis, assessing the impact of individual studies and outliers on the overall results, demonstrated that no single study exerted a significant impact on the overall pooled estimate (Table 3).

**Table 3.**
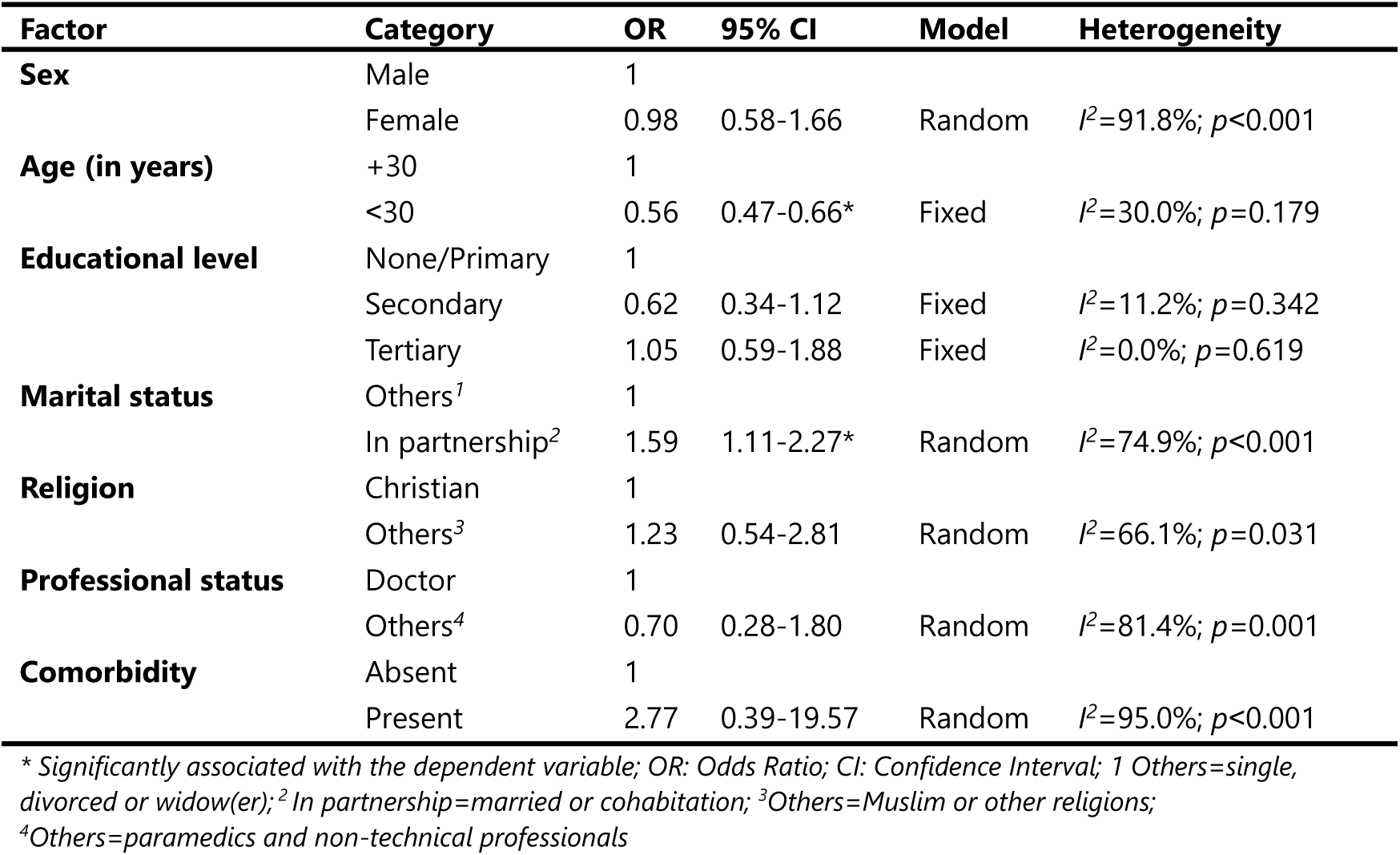
Synthesis of COVID-19 vaccine uptake associated factors in Cameroon, 2021-2024.

### Determinant of Vaccine Uptake

Participant age and marital status were significantly associated with the vaccine uptake. Individuals under 30 years of age were 89% less likely to receive the vaccine compared with those 30 years and older (OR=0.53; 95% CI: 0.42-0.67). People in partnership in the country were 59% more likely to be vaccinated than those in other marital status categories (OR=1.59; 95% CI: 1.11-2.27). Although non statistically significant, the presence of comorbidity showed a strong positive association with COVID-19 vaccine uptake (OR=2.77; 95% CI: 0.39-19.57) (Fig 6, Table 4).

**Fig. 6.**
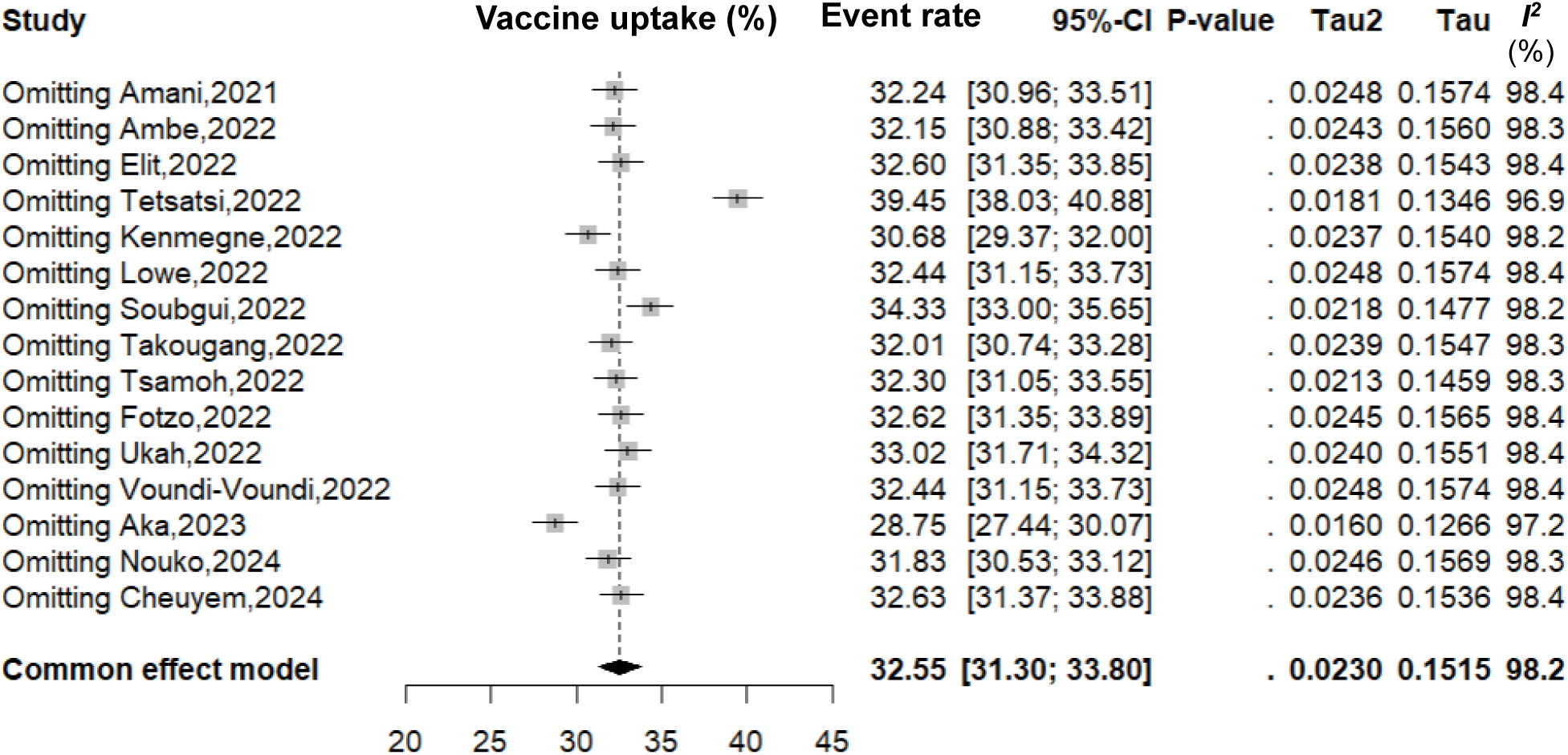
Sensitivity analysis exploring robustness of vaccine coverage pooled estimate, 2021-2024

**Fig. 7.**
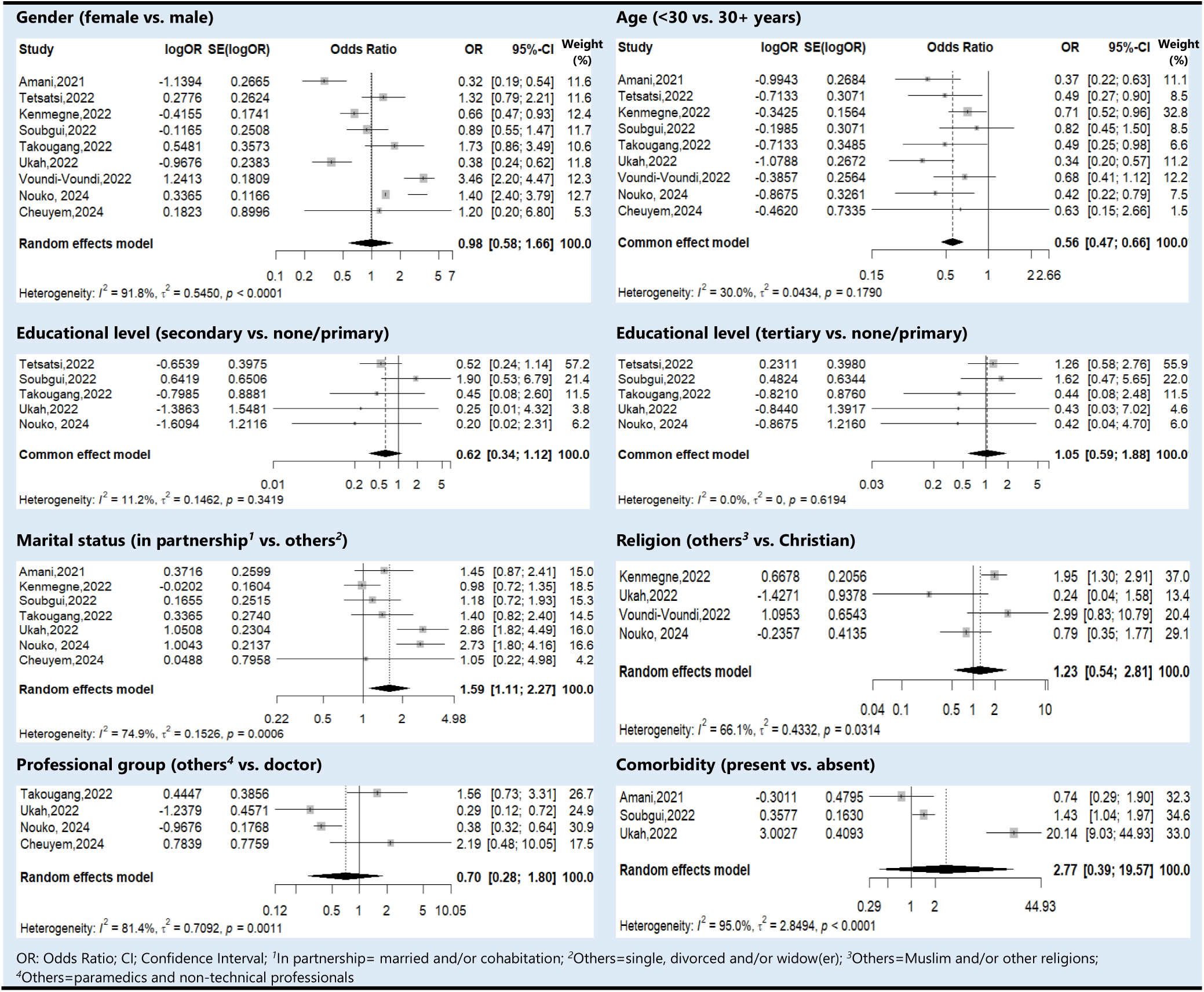
Pooled estimates of the strength of some COVID-19 vaccine uptake determinants in Cameroon, 2021-2024

**Table 4.**
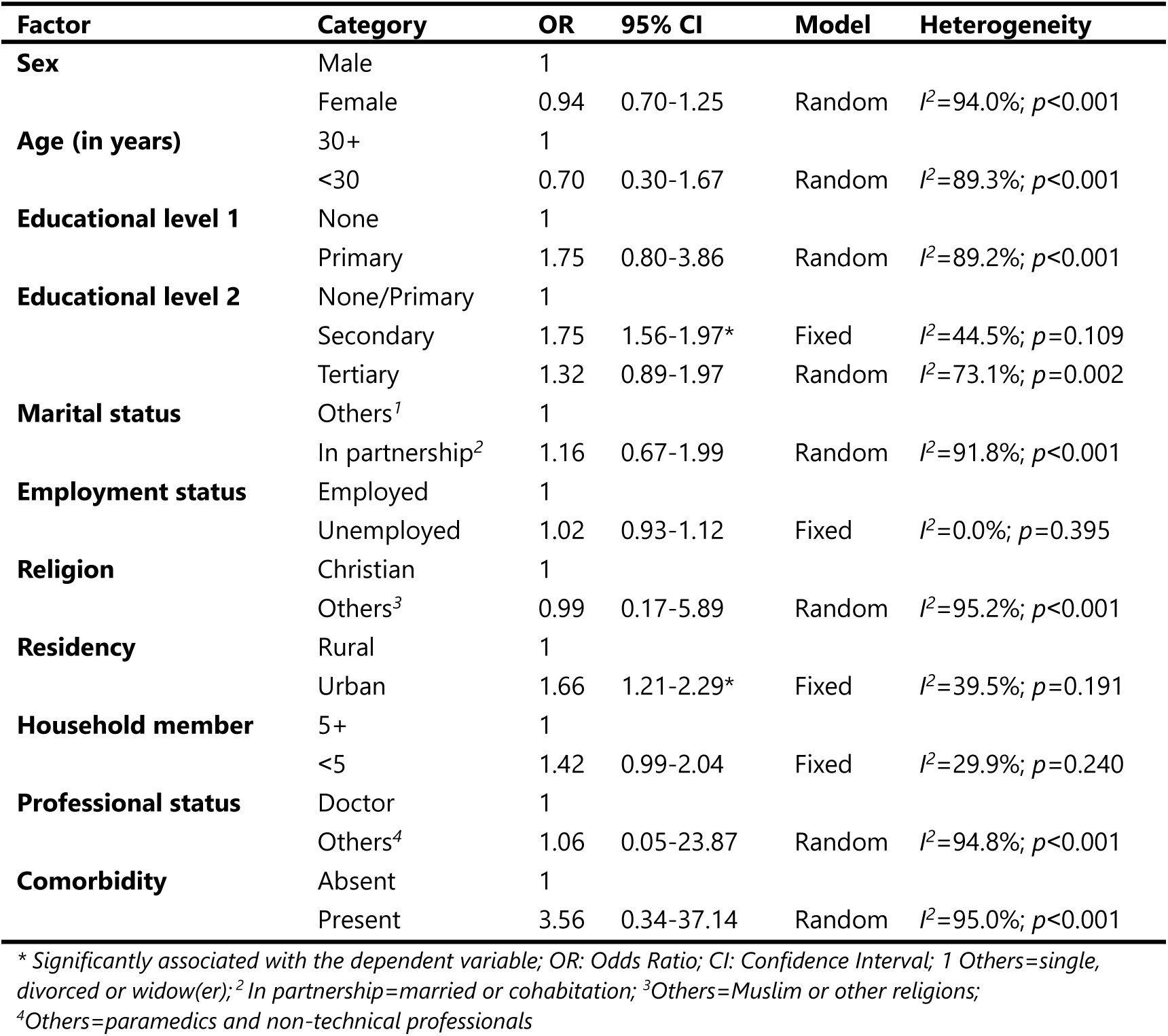
Synthesis of factors associated with COVID-19 vaccine acceptance in Cameroon, 2021-2024.

### Determinant of Vaccine Acceptance

People with a secondary level of education were significantly 75% more willing to accept the COVID-19 vaccine than those with no formal education or a primary level of education (OR = 1.75; 95% CI: 1.56–1.97). Those residing in urban areas were 1.4 times more likely to accept the vaccine than those residing in rural areas (OR = 1.66; 95% CI: 1.21–2.29). The odds of accepting the vaccine were almost four times higher among individuals living with one or more chronic conditions than those without comorbidity, although this association was not statistically significant (OR = 3.56; 95% CI: 0.34–37.14) (Table 5, Supplemental Figure 1 to 12).

## Discussion

This study was conducted to determine the proportion of vaccine uptake and identify associated factors. A total of 22 reports were retrieved from online platforms and gray literature. The inclusion of gray literature enabled the incorporation of unpublished findings and reduced the potential for publication bias arising from the non-publication of negative results.

### Vaccination Coverage

The overall pooled vaccination coverage was 37.14% (95% CI: 29.24-45.05) exhibiting a high level of heterogeneity. This vaccination coverage was significantly lower than the 70% target set by global health authorities for the end of 2022 [9]. Our study demonstrates relatively stable vaccination coverage across study (30-40%) across the study period (2021-2024) reflecting persistent vaccine barriers (including hesitancy) and mitigate effect of effort to increase vaccine uptake. The country has conducted several round of vaccination against the COVID-19 since the vaccination campaign launch in March 2021, with the most recent occurring in November 2022 [7]. Despite national effort to achieve herd immunity within community by reaching at least 60% vaccination coverage, vaccine hesitancy remained of critical obstacle to achieving this target [6,12]. Studies have identified various barriers to vaccination, including concerns about vaccine-related adverse, doubts about the composition of the vaccine, and blood-injection-injury fears which may explain approximately 10% of cases of COVID-19 vaccine hesitancy [10,37].

A metanalysis from Ethiopia in 2024 reported a pooled vaccination coverage (29.6%; 95% CI: 28.7-30.6), a finding not significantly different from our results. Our findings corroborate observations from a weekly report of April 2024, which indicated that 38% of the African Region’s population had received ≥1 dose of COVID-19 vaccine [14].

Regarding geographical trends, with the exception of the North-west Region, the lowest pooled vaccination coverage was observed in the South-west Region (35.82 %; 95% CI: 18.91-52.73). They represent Regions where several armed groups steer security challenges since almost a decade in Cameroon [38,39]. Studies have highlighted the adverse impact of the crisis on the health system in general and on the immunization service in particular, which could explain the observed COVID-19 vaccination coverage [38–40]

Healthcare workforce is one of the six pillars of the healthcare system [41,42]. Their contribution to the fight against the pandemic was essential during the implementation of the response strategy and has been acknowledged by global health authorities [43,44]. This population group presented the highest pooled estimate of vaccine uptake in Cameroon (42.12%; 95% CI: 34.14-50.09). This is significant, as they in forefront during pandemic management and their compliance with vaccination is critical not only to prevent their own infection by this life-threatening disease but also to limit the disease transmission whiting healthcare settings [44,45]. Our findings, however, were significantly lower than the reported vaccine uptake among healthcare workers in Africa (65.6%) and worldwide (77.3%) [13]. This suggest that efforts are still needed to improve vaccination coverage among this professional group, which is particularly vulnerable during pandemics.

### Vaccination Determinant

Individuals under 30 years of age were 89% significantly less likely to receive the vaccine compared with those aged 30 years and older. Several factor could explain these results. The Risk perception may be lower among younger adults who generally perceive themselves as being at lower risk of severe COVID-19 illness compared to older adults. This lower perceived risk can translate to a decreased perceived need for vaccination. Misinformation and social media influence may have a greater impact among younger adults who are often more active on social media platforms where misinformation and vaccine hesitancy narratives can proliferate rapidly. This exposure can negatively influence their attitudes towards vaccination. The prioritization of older age groups and those with comorbidities during initial vaccine rollouts in many countries may have contributed to a perception among younger adults that vaccination was less urgent for them. A meta-analysis in Ethiopia found that younger adult exhibited lower acceptance of the COVID-19 vaccine [46].

People in partnership in the country were 59% more likely to be vaccinated than those in other marital status categories. Shared decision-making and social influence within the household may explain this finding. In fact, individuals in partnerships may discuss health decisions, including vaccination, with their partners. This shared decision-making process could lead to increased vaccine uptake if one partner is inclined to be vaccinated. Furthermore, if one partner is vaccinated, the other may be more likely to follow suit due to concerns about protecting their partner and maintaining a healthy household.

Although not statistically significant, the presence of comorbidity showed a strong positive association with COVID-19 vaccine uptake (OR=2.77; 95% CI: 0.39-19.57). This observation aligns with the understanding that people with underlying health conditions are at higher risk of severe COVID-19, making vaccination more important for them. Further research with larger sample sizes is warranted to confirm this association.

## Strength and Limitations

The use of local unpublish research data (gray literature) helped to address potential publication bias. The absence of association assessments of vaccine uptake and acceptance in some studies have reduced the number of studies included in pooled odds ratio assessment and should be considered in interpreting the result related to vaccination determinants. However, the choice effects model adopted for each determinant were done according to the heterogeneity index. The use of cross-sectional studies design to assess associated factor limits the ability to establish the temporal relationship between exposure and outcome. The study relied on self-reported data, which may be susceptible to social desirability bias, potentially leading to overreporting of vaccine acceptance or uptake. Because we used crude odds ratios to assess determinants, the observed associations may be influenced by unmeasured confounding factors not accounted for in the analysis, such as socioeconomic status, access to healthcare, and cultural beliefs. The findings should be interpreted in the context of the specific time period and geographical location of the included studies.

## Conclusions

In conclusion, this meta-analysis revealed a suboptimal pooled COVID-19 vaccine uptake proportion of 37.14% in Cameroon from 2021 to 2024, significantly lower than the global target of 70% required to ensure herd immunity. Subgroup analyses identified lower uptake among the general population and in community settings, whereas healthcare workers demonstrated the highest uptake. Geographically, the two Regions experiencing armed conflicts (North-west and South-west Regions) exhibited the lowest uptake. Factors such as younger age and not being in a partnership were significantly associated with lower vaccine uptake. Besides, the level of education and area of residency were significantly associate with vaccine acceptance. These findings underscore the crucial need to scale up efforts to improve vaccination coverage, particularly among the most vulnerable populations. Public health interventions should be tailored to address the specific concerns and needs of different age groups and marital statuses. These findings may also inform decision-making during response to future emerging and re-emerging disease epidemics.

## Data Availability

All data generated or analyzed during this study are included in this published article and supplemental material.

## Abbreviations

CI: Confidence Interval
COVID-19: New Coronavirus Disease
CS: Cross Sectional Study
HCW: Healthcare worker
MeSH: Medical Subject Headings
OR: Odds Ratio
PRISMA: Preferred Reporting Items for Systematic Reviews and Meta-Analysis
VA: Vaccine Acceptance
VU: Vaccine Uptake

## Declarations

### Author contributions

FZLC conceived the original idea of the study. FZLC and CA conducted the literature search. FZLC, CA and CD selected the studies, extracted the relevant information, and synthesized the data. FZLC performed the analyses and wrote the first draft of the manuscript. All authors critically reviewed and revised successive drafts of the manuscript. All authors read and approved the final manuscript.

### Ethical Approval Statement

Not applicable

### Consent for publication

Not applicable.

### Availability of data and materials

Source of data supporting this systematic review are available in the reference. All data generated or analyzed during this study are included in this published article and supplemental material.

### Competing interests

All authors declare no conflicts of interest and have approved the final version of the article.

### Funding source

This research did not receive any specific grant from funding agencies in the public, commercial or not-for-profit sectors.

## Notes

### Competing Interest Statement

The authors have declared no competing interest.

### Author Declarations

All data source are available in the reference list.

## References

1. Mengistu DA, Demmu YM, Asefa YA. Global COVID-19 vaccine acceptance rate: Systematic review and meta-analysis. Front Public Health. 2022; 10:1044193.

2. Tchiasso D, Mendjime P, Fai KN, Wandji BSN, Yuya F, Youm É, et al. Dynamic factors associated with COVID-19 vaccine uptake in Cameroon between 2021 and 2022. J Public Health Afr. 2024;15(1):8.

3. Shattock AJ, Johnson HC, Sim SY, Carter A, Lambach P, Hutubessy RCW, et al. Contribution of vaccination to improved survival and health: modelling 50 years of the Expanded Programme on Immunization. The Lancet. 2024;403(10441):2307–16.

4. Ioannidis JPA, Pezzullo AM, Cristiano A, Boccia S. Global estimates of lives and life-years saved by COVID-19 vaccination during 2020-2024. medRxiv; 2024. doi: 10.1101/2024.11.03.24316673v2

5. Meslé MMI, Brown J, Mook P, Katz MA, Hagan J, Pastore R, et al. Estimated number of lives directly saved by COVID-19 vaccination programmes in the WHO European Region from December, 2020, to March, 2023: a retrospective surveillance study. Lancet Respir Med. 2024;12(9):714–27.

6. Amani A, Mossus T, Cheuyem FZL, Bilounga C, Mikamb P, Basseguin Atchou J, et al. Gender and COVID-19 Vaccine Disparities in Cameroon. COVID. 2022;2(12):1715–30.

7. Amani A, Njoh AA, Mouangue C, Zobel CLF, Mossus T. Vaccination Coverage and Safety in Cameroon; Descriptive Assessment of COVID-19 Infection in Vaccinated Individuals. Health Sci Dis. 2022;23(8):1–8.

8. Aka TK, Atanga SN, Esemu SN, Ndip LMA. Attitudes, and perceptions of COVID-19 vaccines and Acceptance to receive COVID-19 vaccine among healthcare workers in Yaounde, Cameroon. OALib. 2024;11(04):1–23.

9. Cameroon steps up vaccination efforts with support from WHO, UNICEF, WB and other partners. WHO, Geneva. 2022. https://www.who.int/news/item/11-12-2022-cameroon-steps-up-vaccination-efforts-with-support-from-who--unicef--wb-and-other-partners. Accessed 2025 Jan 7.

10. Takougang I, Cheuyem FZL, Lyonga EE, Ndungo JH, Mbopi-Keou FX. Observance of Standard Precautions for Infection Prevention in The Covid-19 Era: A Cross Sectional Study in Six District Hospitals in Yaounde, Cameroon. Am J Biomed Sci Res. 2023;19(5):590–8.

11. Baghani M, Fathalizade F, Loghman AH, Samieefar N, Ghobadinezhad F, Rashedi R, et al. COVID-19 vaccine hesitancy worldwide and its associated factors: a systematic review and meta-analysis. Sci One Health. 2023; 2:100048.

12. Cheuyem FZL, Amani A, Nkodo ICA, Boukeng LBK, Edzamba MF, Nouko A, et al. COVID-19 Vaccine Acceptance and Hesitancy in Cameroon: A Systematic Review and Meta-analysis. medRxiv; 2024. https://www.medrxiv.org/content/10.1101/2024.12.12.24318938v1. Accessed 2024 Dec 13.

13. Galanis P, Vraka I, Katsiroumpa A, Siskou O, Konstantakopoulou O, Katsoulas T, et al. COVID-19 Vaccine Uptake among Healthcare Workers: A Systematic Review and Meta-Analysis. Vaccines. 2022;10(10):1637.

14. Doshi RH. COVID-19 Vaccination Coverage — World Health Organization African Region, 2021–2023. MMWR Morb Mortal Wkly Rep. 2024;73(14):307–11.

15. Moher D, Liberati A, Tetzlaff J, Altman DG, Group TP. Preferred Reporting Items for Systematic Reviews and Meta-Analyses: The PRISMA Statement. PLOS Med. 2009;6(7): e1000097.

16. Cheuyem FZL, Mouangue C, Ajong BN, Edzamba MF, Hamadama DCM, Achangwa C, et al. Occupational Exposures to Blood and other Body Fluids among Healthcare Workers in Cameroon: A Systematic Review and Meta-analysis. medRxiv; 2024. doi: 10.1101/2024.12.05.24318564v2.

17. JBI Critical Appraisal Tools. The Joanna Briggs Institute, Adelaide. 2017. https://jbi.global/critical-appraisal-tools. Accessed: 2024 Dec 3.

18. R Core Team. R: A Language and Environment for Statistical Computing. R Foundation for Statistical Computing, Vienna, Austria. 2024 https://www.R-project.org/. Accessed: 2024 Dec 19.

19. Abongwa LE, Sumo L, Ngum NH, Muhammed NN, Njiwale MS, Nakuh NM, et al. A Survey on Factors Influencing COVID-19 Vaccine Hesitancy in Bamenda-Cameroon. J Adv Microbiol. 2022;1–14.

20. Djuikoue CI, Kamga Wouambo R, Pahane MM, Demanou Fenkeng B, Seugnou Nana C, Djamfa Nzenya J, et al. Epidemiology of the Acceptance of Anti COVID-19 Vaccine in Urban and Rural Settings in Cameroon. Vaccines. 2023;11(3):625.

21. Gunawardhana N, Baecher K, Boutwell A, Pekwarake S, Kifem M, Ngong MG, et al. COVID-19 vaccine acceptance and perceived risk among pregnant and non-pregnant adults in Cameroon, Africa. Sallam M, editor. PLOS ONE. 2022;17(9):e0274541.

22. Ngasa NC, Ngasa SN, Tchouda LAS, Tanisso E, Abanda C, Dingana TN. Spirituality and other factors associated with COVID-19 Vaccine Acceptance amongst Healthcare Workers in Cameroon. Research Square; 2021. doi:10.21203/rs.3.rs-712354/v1.

23. Tambo E, Tsague CL, Ebong SB, Tchuendem I, Ngazoue EF, Fankep B, et al. Acceptability Of Covid-19 Vaccines And Vaccination In Cameroon: Challenges And Way Forward. Research Square; 2022. doi:10.21203/rs.3.rs-1369012/v1.

24. Ambe NC, Akum AE, Binwi NF, Ngunde PJ. Prevalence, perceptions and factors influencing covid-19 vaccines’ uptake among nurses in fako division, cameroon. medRxiv; 2023. doi: 10.1101/2023.01.25.23284999.

25. Dinga JN, Njoh AA, Gamua SD, Muki SE, Titanji VPK. Factors Driving COVID-19 Vaccine Hesitancy in Cameroon and Their Implications for Africa: A Comparison of Two Cross-Sectional Studies Conducted 19 Months Apart in 2020 and 2022. Vaccines. 2022;10(9):1401.

26. Elit L, Ngalla C, Afungchwi G, Tum E, Fokom-Domgue J, Nouvet E. Perceptions of COVID 19 Vaccine in Rural Cameroon. Med Discoveries. 2023;2(1):1007.

27. Fotzo NNC, Takougang I, Nokam A. Needle Stick Injuries: Prevalence and Reporting Patterns Among Bucco Dental And Medical Health Workers In Two Reference Hospitals In Yaoundé. Thesis. Yaoundé: Faculty of Medicine and Biomedical Sciences, The University of Yaoundé I; 2022.

28. Christelle KNE, Ngha KJ, Regine EE, Olga B. Factors Associated with Covid-19 Vaccines Acceptance Among Health Care Professionals in the West Region of Cameroon. Health Res Afr. 2024;2(8):35–42.

29. Lowe JM, Nokam ME, Nseme E, Voundi EV, Songue E, Voundi JV, et al. COVID-19: Comparison of Vaccination Coverage between Oral Health Care Practitioners and Other Health Care Personnel in Yaounde. Immunomforschung;18(2).

30. Moguem Soubgui AF, Foko LPK, Embolo Enyegue EL, Mboussi WSN, Koanga Mogtomo ML. Prevalence, Clinical Profile and Determinants of COVID-19 Vaccination and SARS-CoV-2 Breakthrough Infection in Douala, Cameroon. Trends Med Res. 2023;18(1):122–35.

31. Tetsatsi ACM, Nguena AA, Deutou AL, Talom AT, Metchum BT, Tiotsia AT, et al. Factors Associated with COVID-19 Vaccine Refusal: A Community-Based Study in the Menoua Division in Cameroon. Trop Med Infect Dis. 2023;8(9):424.

32. Tsamoh FF, Takougang I. Organisational determinants of the observance of infection prevention measures within bucco-dental health services in the Mifi and Dschang Health Districts. Thesis. Yaoundé: Faculty of Medicine and Biomedical Sciences, The University of Yaounde 1; 2022.

33. Ukah EC, Tambe J, Tanue AE, Ngeha NC, Shei MC, Tabe OBV, et al. COVID-19 vaccine uptake among healthcare workers in the Limbe Health district of Cameroon. J Public Health Epidemiol. 2024;16(1):28–40.

34. Voundi-Voundi E, Songue E, Voundi-Voundi J, Nseme EG, Abba-Kabir H, Kamgno J. Factors Associated with COVID-19 Vaccine Hesitancy Among Health Personnel in Yaounde, Cameroon. Health Sci Dis. 2023;24(2 Suppl 1):23–7.

35. Cheuyem FZL, Takougang I, Lyonga EE, Kwabong E. Compliance with standard precautions and microbial profile of media in the gynecology-obstetrics department of the University Teaching Hospital of Yaoundé. Thesis. Yaoundé: Faculty of Medicine and Biomedical Sciences, The University of Yaounde I; 2024.

36. Nouko A, Takougang I, Nguefack F. Determinants of compliance to vaccination as a means of infection prevention among healthcare workers in the health districts of Yaoundé Thesis. Yaoundé: Faculty of Medicine and Biomedical Sciences, The University of Yaounde I; 2024.

37. Freeman D, Lambe S, Yu LM, Freeman J, Chadwick A, Vaccari C, et al. Injection fears and COVID-19 vaccine hesitancy. Psychol Med. 2023 Mar;53(4):1185–95.

38. Njoh AA, Saidu Y, Bachir HB, Ndoula ST, Mboke E, Nembot R, et al. Impact of periodic intensification of routine immunization within an armed conflict setting and COVID-19 outbreak in Cameroon in 2020. Confl Health. 2022;16(1):29.

39. Takougang I, Cheuyem FZL, Changeh BA, Nyonga ND, Moneboulou HM. Accidental Exposure to Body Fluids Among Healthcare Workers in a Referral Hospital in the Security-Challenged Region of South West Cameroon. J Nurs Healthc. 2024;9(2):1–13.

40. Saidu Y, Vouking M, Njoh AA, Bachire HB, Tonga C, Mofor R, et al. The effect of the ongoing civil strife on key immunisation outcomes in the North West and South West regions of Cameroon. Confl Health. 2021;15(1):8.

41. Cheuyem FZL, Ajong BN, Amani A, Boukeng LBK, Ngos CS, Nkongo FK, et al. Sustainable Development Goals and Health Sector Strategic Indicators Assessment in Cameroon: A Retrospective Analysis at Regional and National Levels. medRxiv; 2024. doi: 10.1101/2024.11.25.24317921v1

42. Cheuyem FZL, Amani A, Ajong BN, Boukeng LBK, Mouangue C, Tsafack MGM, et al. Humanization of Care: A Geospatial Analysis of Key Indicators of Quality and Safety of Health Care and Service in the Centre Region, Cameroon. medRxiv; 2024 doi: 10.1101/2024.11.11.24317125v1.

43. Almaghrabi RH, Alfaradi H, Hebshi WAA, Albaadani MM. Healthcare workers experience in dealing with Coronavirus (COVID-19) pandemic. Saudi Med J. 2020;41(6):657–60.

44. Chemali S, Mari-Sáez A, El Bcheraoui C, Weishaar H. Health care workers’ experiences during the COVID-19 pandemic: a scoping review. Hum Resour Health. 2022;20(1):27.

45. Takougang I, Lekeumo Cheuyem FZ, Ze BRS, Tsamoh FF, Moneboulou HM. Awareness of standard precautions, circumstances of occurrence and management of occupational exposures to body fluids among healthcare workers in a regional level referral hospital (Bertoua, Cameroon). BMC Health Serv Res. 2024;24(1):424.

46. Tolossa T, Fetensa G, Feyisa BR, Wakuma B, Lema M. Willingness to accept COVID-19 vaccine and its determinants in Ethiopia: A systematic review and meta-analysis. Front Virol. 2023; 3:1065991.

