## Supplemental file for "COVID-19 Vaccine Uptake and its Determinants in Cameroon: A Systematic Review and Meta-analysis"

**Determinant of COVID-19 vaccine acceptance**

<u>Sex (female vs male)</u>

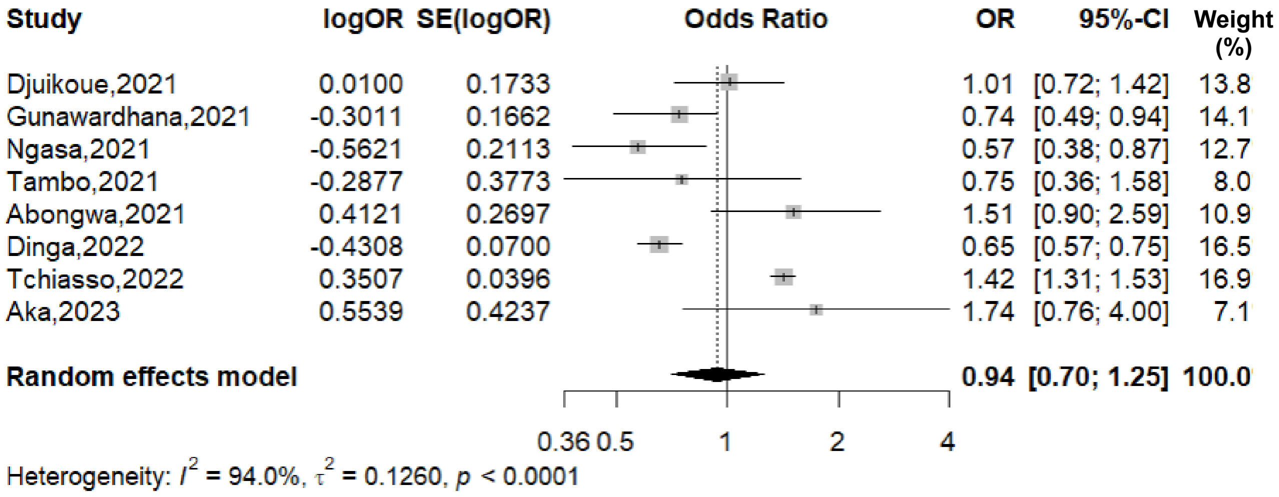


**Fig. 1** Association between the sex and the COVID-19 vaccine acceptance in Cameroon, 2021-2023

<u>Age (<30 vs 30+ years)</u>

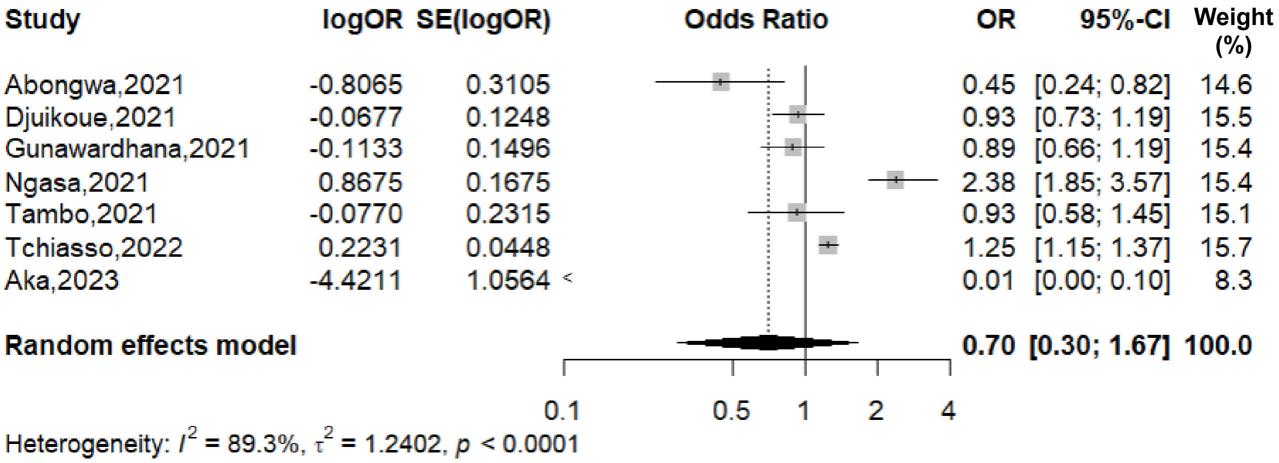


**Fig. 2** Association between the age and the COVID-19 vaccine acceptance in Cameroon, 2021-2022

Educational level (primary vs none)

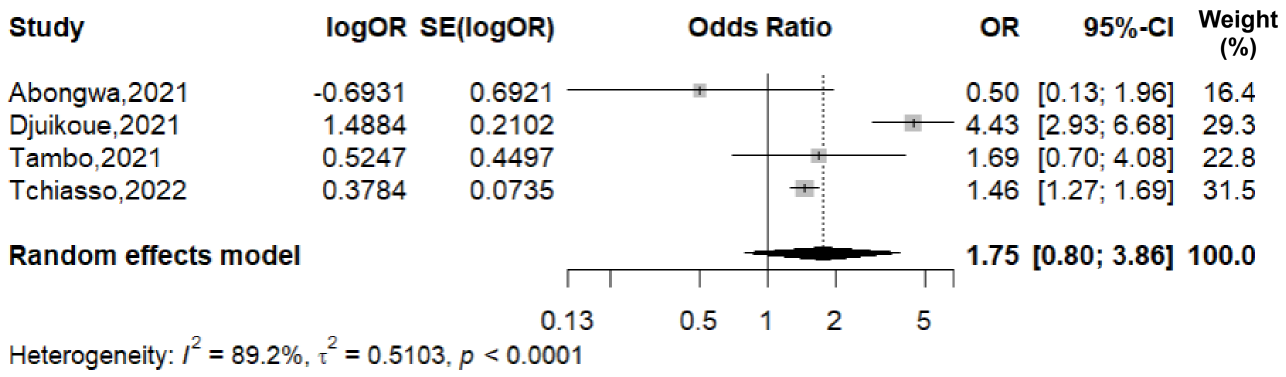


**Fig. 3** Association between the educational level and the COVID-19 vaccine acceptance in Cameroon, 2021-2022

Educational level (secondary vs none/primary)

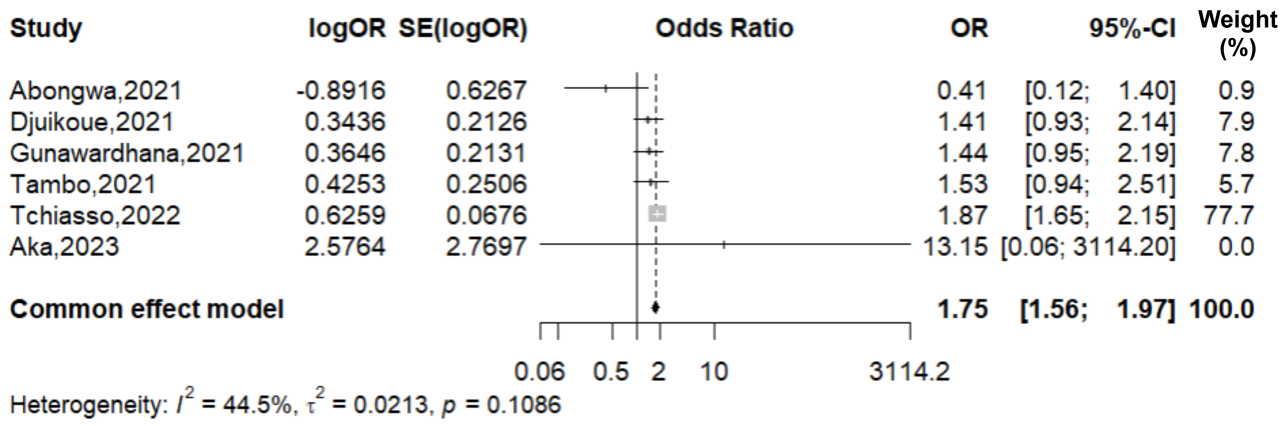


**Fig. 4** Association between the educational level and the COVID-19 vaccine acceptance in Cameroon, 2021-2023

Educational level (tertiary vs none/primary)

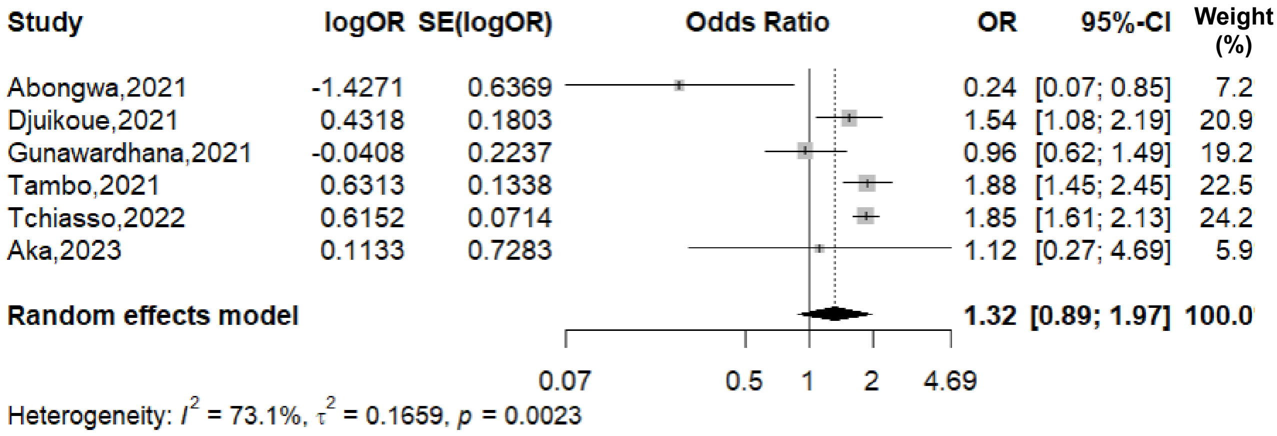


**Fig. 5** Association between the educational level and the COVID-19 vaccine acceptance in Cameroon, 2021-2023

Marital status (in partnership vs others)

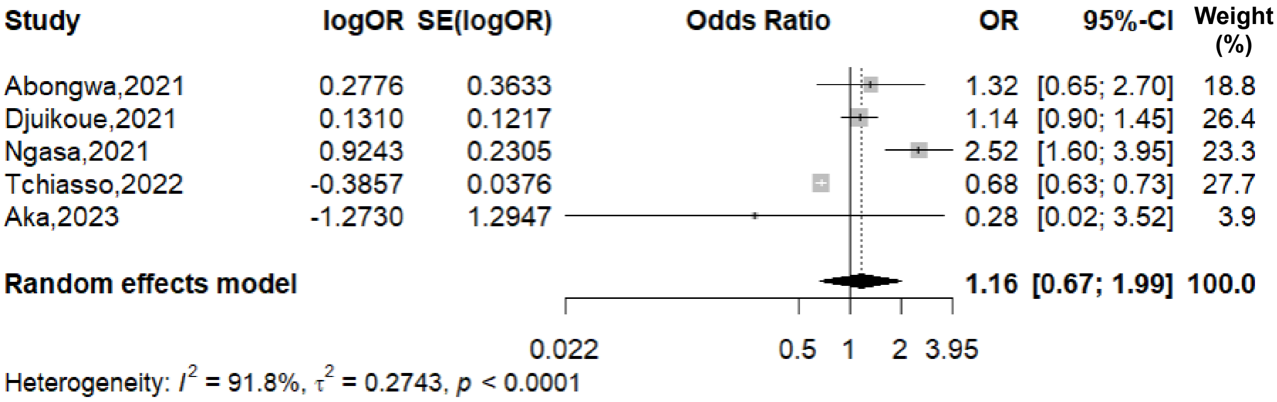


**Fig. 6** Association between the marital status and the COVID-19 vaccine acceptance in Cameroon, 2021-2023

Employment status (unemployed vs employed)

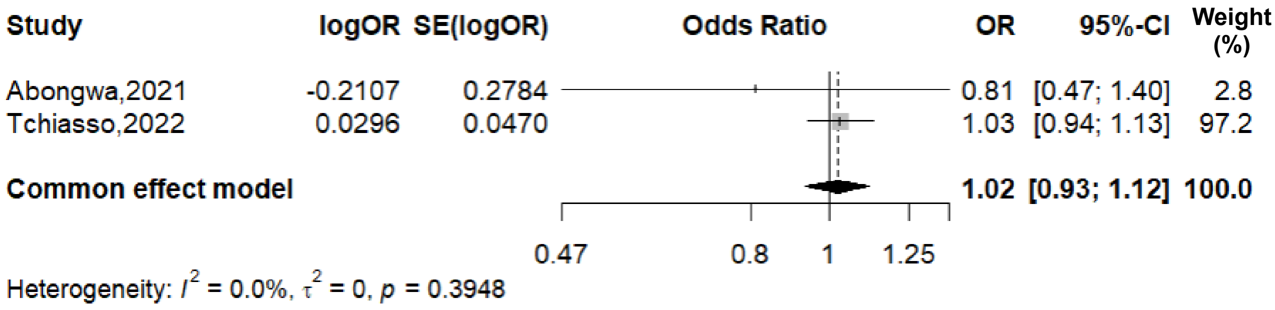

**Fig. 7** Association between the employment status and the COVID-19 vaccine acceptance in Cameroon, 2021-2022

Religion (other vs christian)

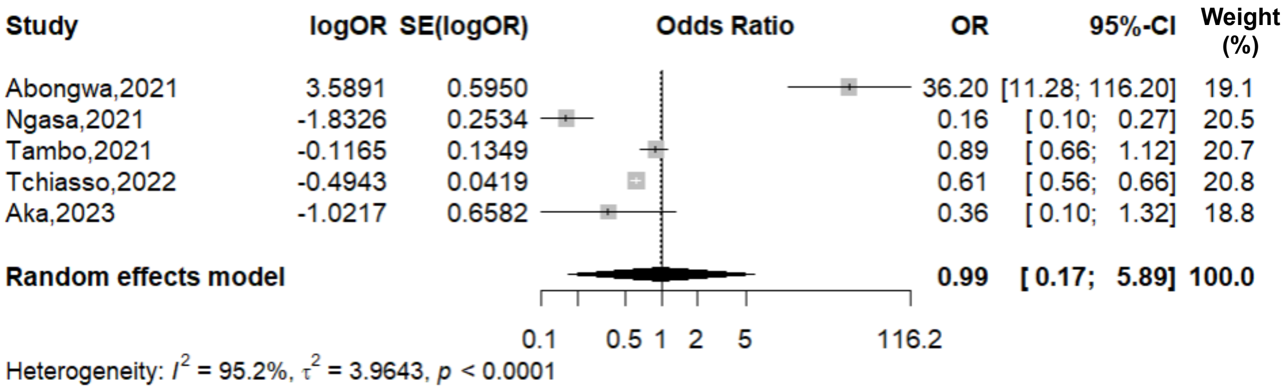


**Fig. 8** Association between the religion and the COVID-19 vaccine acceptance in Cameroon, 2021-2023

Residency (urban vs rural)

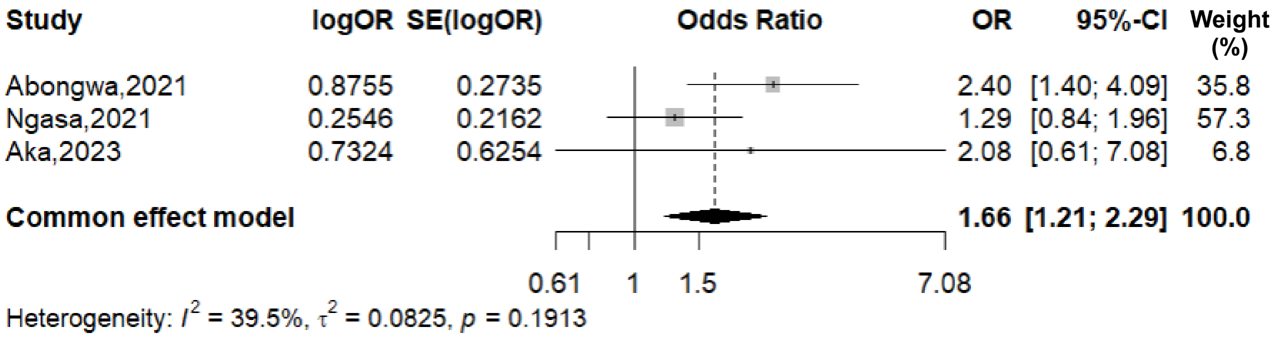


**Fig. 9** Association between the place of residence and the COVID-19 vaccine acceptance in Cameroon, 2021-2023

Professional status (others vs doctor)

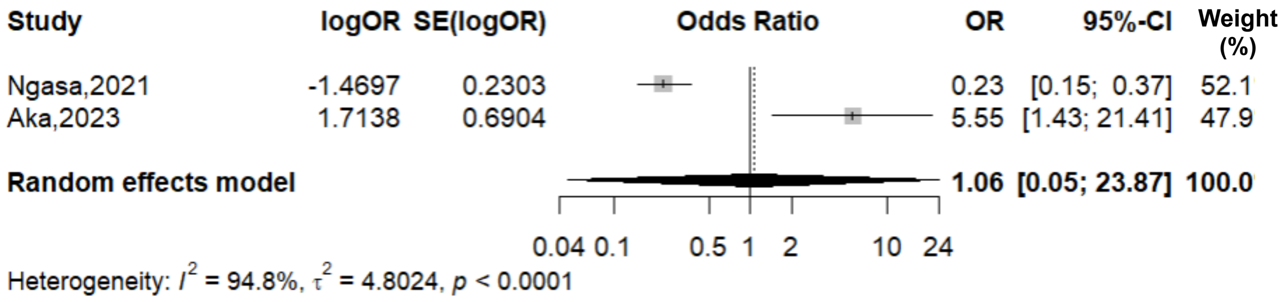


**Fig. 10** Association between the professional status and the COVID-19 vaccine acceptance in Cameroon, 2021-2022

Comorbidity (present vs absent)

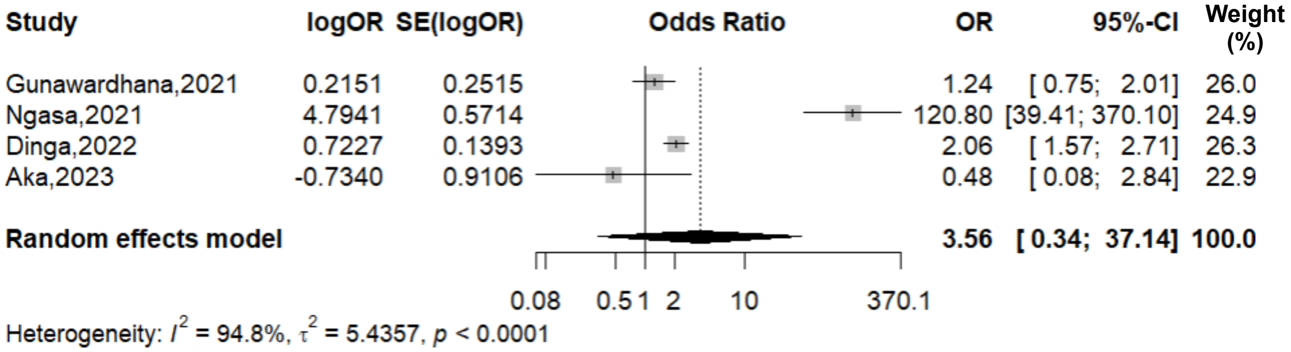


**Fig. 11** Association between the presence comorbidity and the COVID-19 vaccine acceptance in Cameroon, 2021-2023

Number of household members (<5 vs 5+)

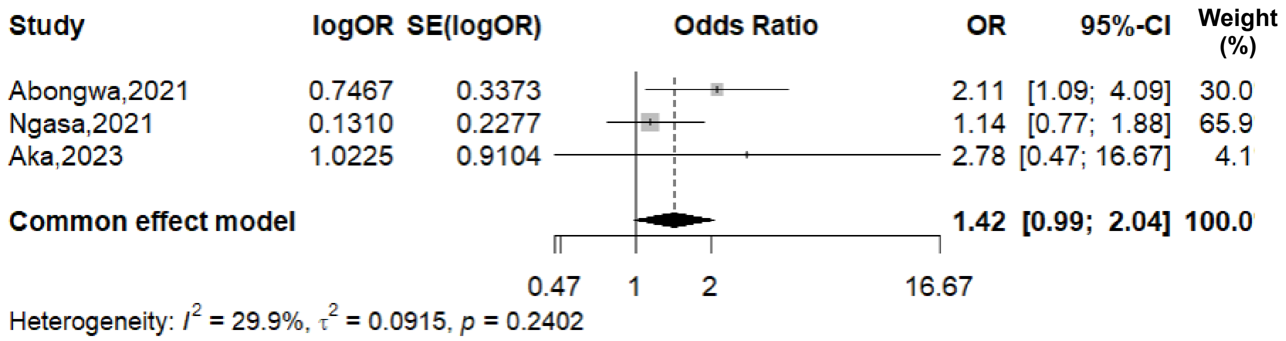


**Fig. 12** Association between the number of household members and the COVID-19 vaccine acceptance in Cameroon, 2021-2022
